# Finding Safety Signals in Sparse Data: Zero-Inflated Models for Pharmacovigilance studies

**DOI:** 10.64898/2026.09.21.26363547

**Authors:** Martin Man, Vicky Cheng, Anthony R. Cox, Christina L. Easter, Alan M. Jones

**Affiliations:** School of Pharmacy, School of Health Sciences, College of Medicine and Health, University of Birmingham, Edgbaston, Birmingham B15 2TT, UK; Department of Applied Health Sciences, School of Health Sciences, College of Medicine and Health, University of Birmingham, Birmingham B15 2TT, UK

**Keywords:** Pharmacovigilance, zero-inflated, ZINB, disproportionality, statistics, SSRI, MHRA

## Abstract

Signal detection is a core task in pharmacovigilance, and disproportionality analysis (DA) remains the dominant approach for spontaneous reporting systems (SRS). However, SRS data are sparse and heavily zero-inflated, reflecting rare adverse drug reactions (ADRs) and substantial under-reporting, which can distort standard count-model assumptions. Using Medicines and Healthcare products Regulatory Agency (MHRA) interactive Drug Analysis Profiles (iDAPs) for selective serotonin reuptake inhibitors (SSRIs), we derived system organ class (SOC)–level adverse event counts. Goodness-of-fit testing indicated that SOC counts were variably consistent with Poisson or negative binomial (NB) distributions, motivating the use of zero-inflated Poisson and zero-inflated NB models. Model adequacy was assessed using Pearson-residual dispersion (absolute fit) and Akaike’s Information Criterion (relative fit). Zero-inflated models provided improved fit for 10 out of all 27 SOCs (*Endocrine disorders, Immune system disorders, Metabolism and nutrition disorders, Infections and infestations, Renal and urinary disorders, Hepatobiliary disorders, Ear and labyrinth disorders, Pregnancy, puerperium and perinatal conditions, Congenital, familial and genetic disorders, Neoplasms benign, malignant and unspecified (including cysts and polyps*)) and yielded signals broadly concordant with DA, including for metabolism-related events. These results support zero-inflated modelling as a complementary signal detection method for iDAPs-derived count data, particularly where internal model-based validation and explicit handling of excess zeros are desired.

**Key Points:**

- Standard disproportionality methods can struggle with sparse pharmacovigilance data because many drug–event combinations have lots of zeros.
- Zero-inflated models better reflected the structure of the data for many system organ classes, improving model fit without changing the overall safety message.
- The approach produced signals broadly consistent with traditional analysis, supporting its use as a complementary method rather than a replacement.
- Handling excess zeros directly may improve detection of rare or under-reported adverse drug reactions in spontaneous reporting systems.
- These findings could strengthen post-marketing safety monitoring for SSRIs and other medicines with sparse reporting patterns.

**Plain Language Summary:** We studied whether a different way of analysing reports of side effects could help spot safety concerns for antidepressants called selective serotonin reuptake inhibitors (SSRIs). Using UK medicines safety reports, we compared a standard counting method with a method designed for data that contain many categories with no reports at all. The new method fitted the data better for many body-system groups and gave signals that were broadly similar to the standard approach, while also handling the large number of empty categories more realistically. This matters because side effects in safety databases are often rare and under-reported, which can make them hard to detect. Our findings suggest that these newer statistical methods could improve how researchers and regulators monitor medicine safety, especially when the data are sparse and unevenly reported.

## Introduction

Major depressive disorder (MDD) is a growing global health concern. By 2050, the global prevalence of MDD among adults aged 60 years and older is projected to reach 97.04 million cases, representing an age-standardised prevalence rate of 4.5% across 204 countries and territories [1]. In the United Kingdom (UK), antidepressant prescribing has increased by 3.94% since 2023/24 [2]; the most prescribed class of antidepressants are the selective serotonin reuptake inhibitors (SSRIs) [3]. Although SSRIs are a first-line treatment due to improved safety profiles than previous antidepressants [4], the safety landscape of SSRIs requires further investigation [5,6].

The most widely accepted causative mechanism for MDD is the serotonin theory of depression [7]. Due to the traction of this theory, SSRIs were developed whilst assuming its validity [7,8]. However, literature which discusses against its validity has emerged, thus reflecting the gaps in the understanding of the causative mechanism of the disease [7,8].

Given that SSRIs were developed whilst assuming the serotonin theory of depression to be true [7], of which remains as an area of discussion. Insufficient understanding of the causative mechanism of MDD would yield further questions regarding the long-term safety landscape of SSRIs. As such, the widespread use of SSRIs in the population would emphasize the importance post-market surveillance for future adverse drug reactions (ADRs).

In practice, signal detection methods (SDMs) are applied with the aim of obtaining signals from spontaneous reporting systems (SRSs), of which disproportionality analysis (DA) remains as the most common SDM. The produced signals serve as hypotheses for potential further investigation into a suspected causal relationship between a drug and an ADR [9]. A summary of commonly employed SDMs by country is shown in Supplementary Information (S1).

When DA is conducted, signals are identified by comparing reporting proportions of a particular drug-ADR combination [10]. It is important to note that this does not afford risk quantification [9,10]. Traditional DA is performed by comparing the reporting proportion of one drug-ADR combination with all other drugs present in the database [11,12], whereas restricted DA is performed when a sub-group of the dataset is analysed, such as by therapeutic class or gender [9]. Traditional DA is recommended as the first step, whereas restricted DA serves as a subsequent analysis to reduce noise [9].

However, DA does not address inherent limitations such as underreporting, thus, delaying the detection and identification of the safety profile of a drug. Underreporting is explicitly acknowledged as an unavoidable and structural limitation of SRSs [13,14] and its influence on PV studies is regarded as intractable. Alongside DA, logistic regression (LR) with or without Firth’s penalisation are also SDMs [15,16]. Firth’s LR (FLR) is a commendable approach due to the direct addressing of rare events [16,17], of which ADRs are an example [18]. A characteristic of a rare event would be the over-presence of zeroes in the response variable (Y). While FLR accepts this characteristic of Y, there is a trade-off where the uncertainty of odds ratios (ORs) would be greater than without the penalisation [19,20]. However, these approaches do not enable the exploration of the potential causative mechanisms for the absence of a reported ADR. Furthermore, given the ubiquity of LR and FLR, the dichotomisation of count variables, such as the number of ADR events, would induce information loss.

We suggest a SDM which circumvents information loss and produces signals where the quantification of underreporting can be achieved. Therefore, we suggest the use of zero-inflated (ZI) models as a potential SDM due to the mixed nature (Equation 1). The use of ZI models in a healthcare context is not new as Fernandez and Vatcheva [21] have fitted ZI negative binomial (NB) models to hospital length of stay data. They concluded that ZINB may be a viable regression model when, (1) the assumption of zeroes originating from two separate mechanisms has been satisfied, and (2) the observed outcome is sampled from a ZINB distribution.

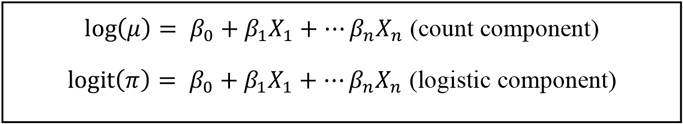

Equation 1: General form of a zero inflated (ZI) model. A ZI model is a mixture model comprised of a count component and a logistic component. The count component models counts from 0 to any integer with respect to the Poisson or negative binomial (NB) distribution, whereas the logistic component models the probability of structural zero (π); it is assumed that there are two distinct zero generating mechanisms which contribute to π [22,23]. In the context of our work, the count component would model the observed ADR counts from 0, whereas the logistic component would model two zero generating mechanisms which contribute to the absence of a report in a spontaneous reporting system (SRS); the absence of a report was either caused by an unreported ADR event which had occurred, or the ADR cannot occur due to a biological or physiochemical cause.

From a methodological standpoint, the rare nature of SSRI-suspected ADRs within SRSs would justify the application of SDMs beyond DA [24]. Furthermore, SSRIs would be chosen in this study given its topicality within public health, the influence of insufficient understanding of the cause of MDD on subsequent development of SSRIs, and the practical justification it presents in favour for the application of ZI models as a candidate SDM. Furthermore, the outputs generated would positively contribute to the greater discussion surrounding the safety concerns of this increasingly prescribed medication [3].

### Aim and objectives

Given that SDMs are applied to SRS data during pharmacovigilance (PV) studies to investigate the safety landscape of any drug, the overall aim of our work is to develop a signal generating workflow (SGW) where the generated signals would be of greater robustness. The present objective was to explore the potential application of ZI models as an SDM. This study explores the operational capability of ZI models as the primary analytical driver in the SGW at the SOC level; DA would be conducted for comparison.

## Methods

### 2.1 Medicines and Healthcare products Regulatory Agency (MHRA)

The MHRA is an executive agency of the Department of Health and Social Care, and alongside marketing authorisation holders (MAH), healthcare professionals (HCPs), patients and the public share responsibility for overseeing PV activities in the UK; the SRS of the MHRA is the Yellow Card Scheme (YCS), which stores all individual case safety reports (ICSR). ICSRs collectively yield an interactive drug analysis profile (iDAP) of each licensed drug. ADRs are recorded per the Medical Dictionary for Regulatory Activities (MedDRA).

### 2.2 Data acquisition

The MHRA disabled data acquisition from the YCS in May 2025 and at time of writing is still pending reinstatement. All iDAP data were downloaded in December 2025 through a legacy hyperlink; all reports were processed up to 19^th^ May 2024. Downloadable content was a folder containing three comma separated value (CSV) files labelled as ‘drug’, ‘case’ and ‘event’, respectively. A full summary of this can be found in Supplementary Information (S2).

### 2.3 System Organ Class (SOC). Abbreviations and in full

### 2.4 Disproportionality analysis (DA)

DA was conducted in R-Studio (version 4.3.1) with the R-package ‘pvda’ (version 0.0.4). DA facilitated by the ‘pvda’ package has been performed previously [25]. A four columned dataset was prepared from the iDAPs, these columns were ‘report ID’, ‘drug’, ‘event’ and ‘group’. The term ‘drug’ contained the names of all drugs in the YCS, ‘event’ contained SOCs, ‘group’ was the gender of the patient (‘Male’, ‘Female’ or ‘Unknown’ if undisclosed).

Reporting odds ratios (RORs), proportional reporting ratios (PRRs) and information components (ICs) were calculated to determine associations between SSRIs and adverse events (AEs). *P*-values were computed separately and corrected with the Benjamini-Hochberg (BH) method. All algorithms, thresholds and method of calculation can be found in Supplementary Information (S3 and S4, respectively). Descriptive statistics for the prepared dataset for DA with the ‘pvda’ package can be found in 3.1 and Supplementary Information (S6).

### 2.5 Pre-analysis prior to ZI model construction

After transformation of the SOC into SOC counts, Goodness of Fit (GoF) tests were performed to determine the distribution of SOC counts, where *p*-value > 0.05 suggested adequate fit to either Poisson or NB distribution. The proportion of zeroes of all SOC counts were also determined. Pre-analysis results for all 27 SOCs are found in the Supplementary Information (S5). Descriptive statistics for predictors included only for regression modelling can be found in 3.3; descriptive statistics for all predictors can be found in Supplementary Information (S7).

### 2.6 Model construction for ZI models

In this study, Poisson and NB models (count models) and ZI models were constructed. Sex-stratified models failed to converge. A summary of the predictors is shown below in Table 2.

**Table 1:** Full list of 27 SOCs of MedDRA. This table summarises how a SOC is abbreviated in MedDRA convention and the full description of each SOC. This report will primarily use the abbreviated format.

| SOC (abbreviation) | SOC (full description) |
| --- | --- |
| Blood | Blood and lymphatic system disorders |
| Card | Cardiac disorders |
| Cong | Congenital, familial and genetic disorders |
| Ear | Ear and labyrinth disorders |
| Endo | Endocrine disorders |
| Eye | Eye disorders |
| Gastr | Gastrointestinal disorders |
| Genrl | General disorders and administration site conditions |
| Hepat | Hepatobiliary disorders |
| Immun | Immune system disorders |
| Infec | Infections and infestations |
| Inj&P | Injury, poisoning and procedural complications |
| Inv | Investigations |
| Metab | Metabolism and nutrition disorders |
| Musc | Musculoskeletal and connective tissue disorders |
| Neopl | Neoplasms benign, malignant and unspecified (incl cysts and polyps) |
| Nerv | Nervous system disorders |
| Preg | Pregnancy, puerperium and perinatal conditions |
| Prod | Product issues |
| Psych | Psychiatric disorders |
| Renal | Renal and urinary disorders |
| Resp | Respiratory, thoracic and mediastinal disorders |
| Repro | Reproductive system and breast disorders |
| Skin | Skin and subcutaneous tissue disorders |
| SocCi | Social circumstances |
| Surg | Surgical and medical procedures |
| Vasc | Vascular disorders |

**Table 2:**
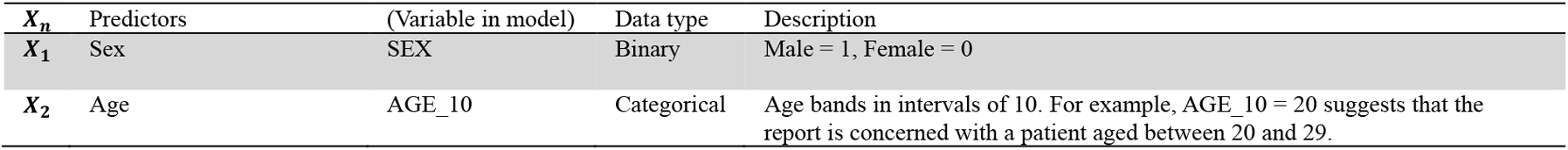
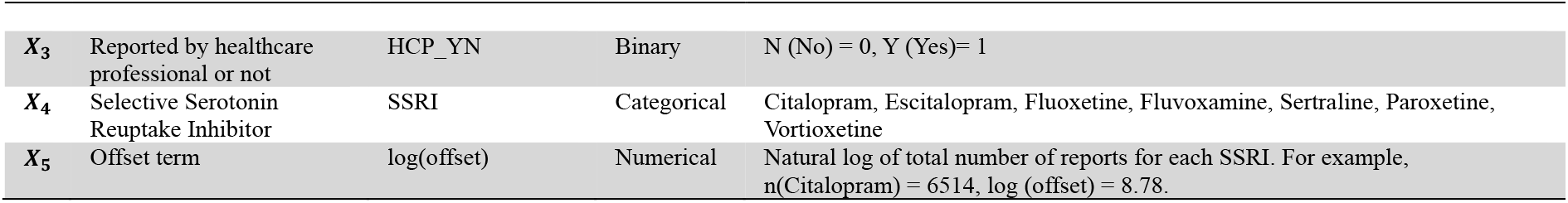
Predictors for Poisson, NB, ZIP and ZINB models.

| $X_n$ | Predictors | (Variable in model) | Data type | Description |
| --- | --- | --- | --- | --- |
| $X_1$ | Sex | SEX | Binary | Male = 1, Female = 0 |
| $X_2$ | Age | AGE_10 | Categorical | Age bands in intervals of 10. For example, AGE_10 = 20 suggests that the report is concerned with a patient aged between 20 and 29. |
| $X_3$ | Reported by healthcare professional or not | HCP_YN | Binary | N (No) = 0, Y (Yes)= 1 |
| $X_4$ | Selective Serotonin Reuptake Inhibitor | SSRI | Categorical | Citalopram, Escitalopram, Fluoxetine, Fluvoxamine, Sertraline, Paroxetine, Vortioxetine |
| $X_5$ | Offset term | log(offset) | Numerical | Natural log of total number of reports for each SSRI. For example, $n(\text{Citalopram}) = 6514$, $\log(\text{offset}) = 8.78$. |

**Table 3:**
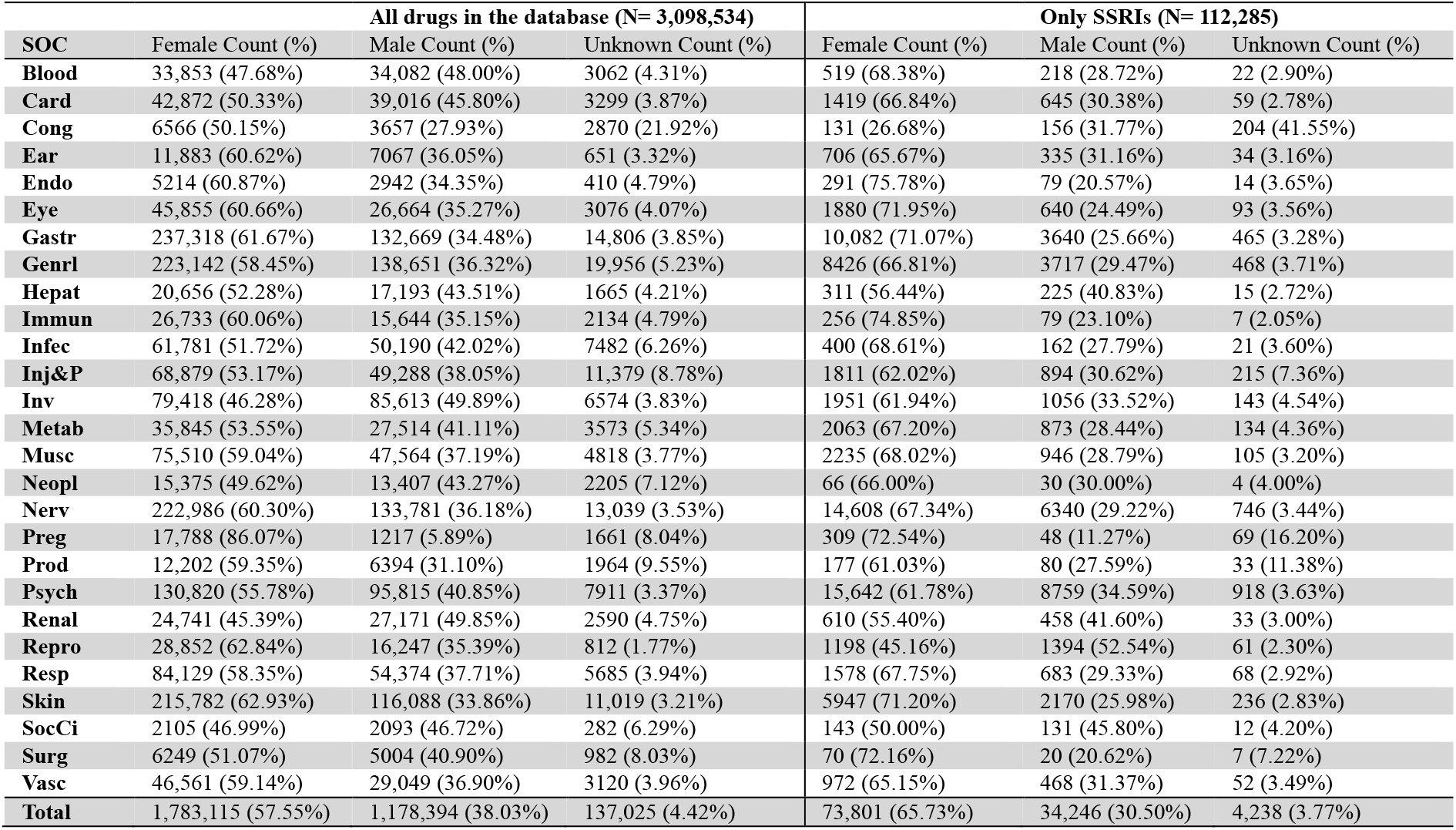
Descriptive statistics for the traditional (left; N= 3,098,534) and restricted (right; N=112,285) DA datasets. Data are presented as count (%) for system organ classes (SOCs) stratified by Female, Male and Unknown.

**Table 4:**
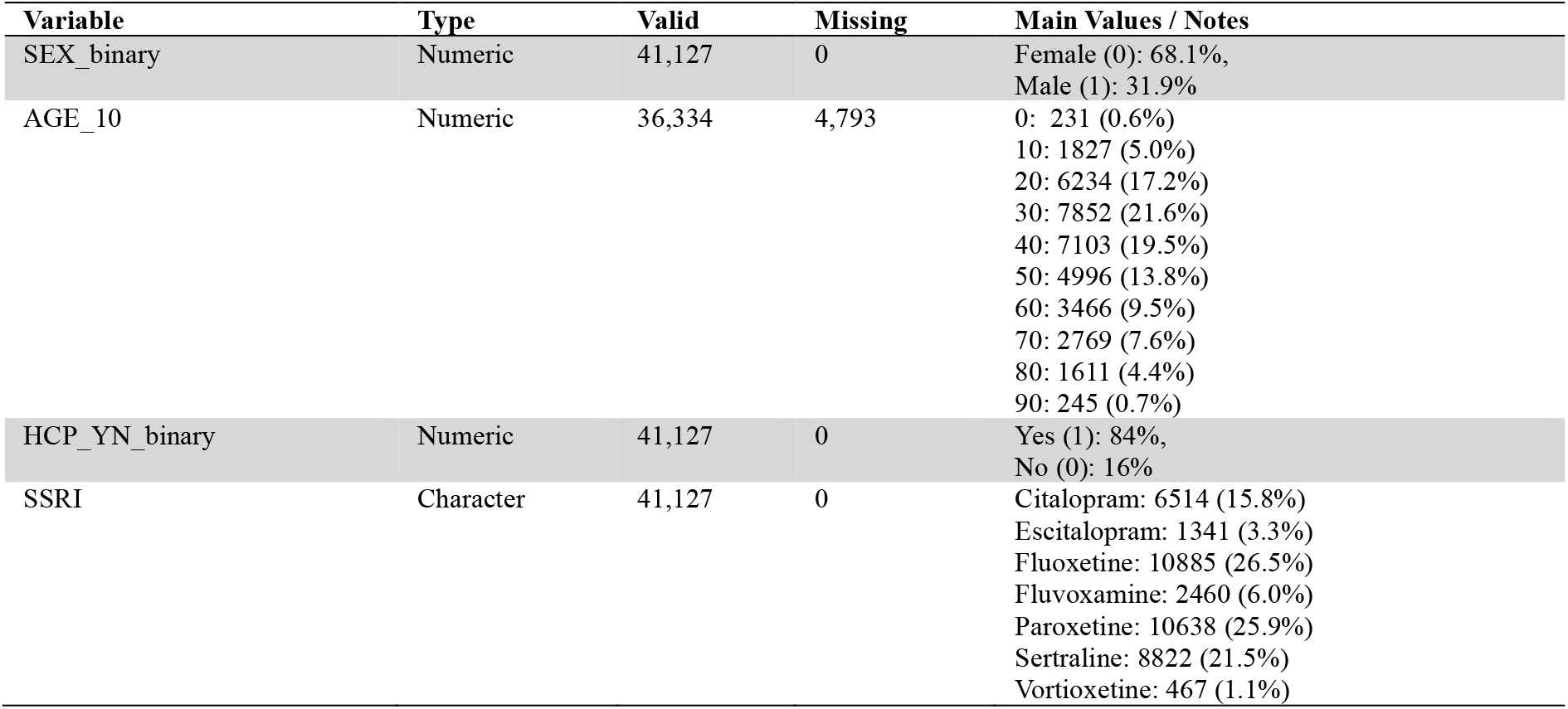
Baseline characteristics of predictors of dataset prepared for fitting of ZI models (N = 41,127). Data type, counts and valid percentages are presented alongside missingness profiles for each variable.

## Results

### 3.1 Descriptive statistics of the prepared dataset for DA

Significant results from DA are shown below. The ‘pvda’ package facilitated the calculation of ROR [95% CI], PRR [95% CI] and IC [IC_025_, IC_0975_]. *P*-values were calculated from formulae presented by Altman and Bland [26]. For a result to be considered significant, all thresholds described in Supplementary Information (S4) had to be met and a Benjamini-Hochberg (BH) corrected *p*-value < 0.05.

### 3.2 Disproportionality analysis results

Prior to applying the BH correction, there were 578 significant signals, 54 remained after correction (45 signals from traditional DA, 9 from restricted DA). While this approach may appear stringent, most disproportionality signals have been regarded as statistical noise [10]. Results of traditional and restricted DA can be subjected to different sources of bias [9]. As such, *p*-value correction increased robustness of signals.

### 3.3 Descriptive statistics for dataset for ZI model fitting

### 3.2 Pre-analysis results

As described in 2.5, GoF tests were conducted, proportion of zeroes and Variance to Mean Ratios (VMRs) were calculated prior to model fitting. These results are shown in Table 5.

**Table 5:**
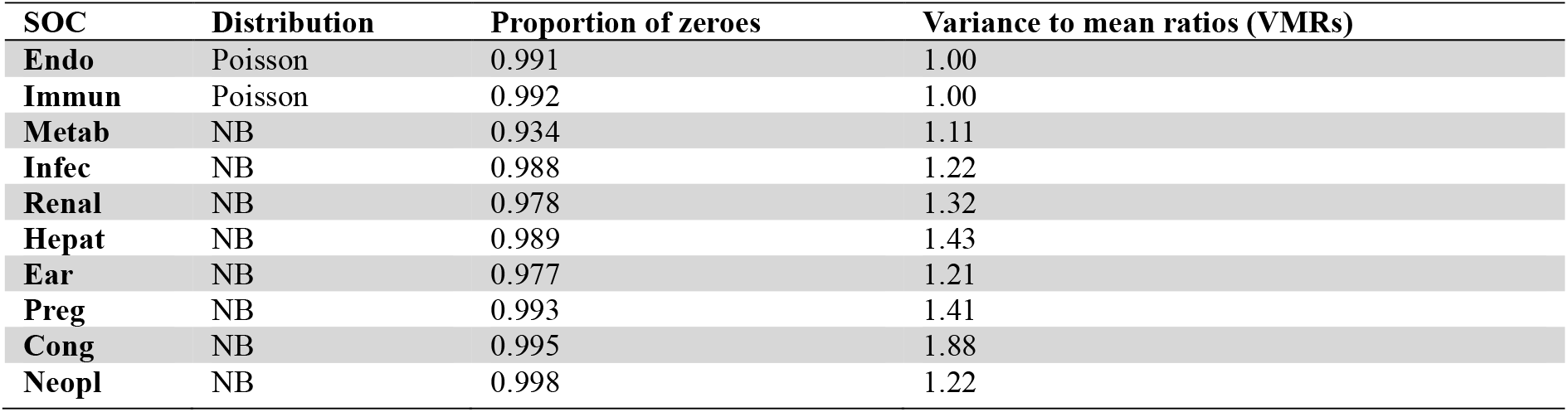
An overview of the pre-analysis results. If the GoF *p*-value was greater than 0.05, the SOC followed a particular distribution. All NB distributed SOCs have VMRs > 1.00, indicating overdispersion, whereas a VMR = 1.00 indicated a lack of overdispersion. Prod and SocCi followed the NB distribution, whereas Surg followed the Poisson distribution. However, these SOCs are not shown nor carried forward for analysis as these SOCs are grounded in human errors. A complete tabulation of the calculated values for all 27 SOCs during the pre-analysis stage can be found in the Supplementary Information (S5).

| SOC | Distribution | Proportion of zeroes | Variance to mean ratios (VMRs) |
| --- | --- | --- | --- |
| Endo | Poisson | 0.991 | 1.00 |
| Immun | Poisson | 0.992 | 1.00 |
| Metab | NB | 0.934 | 1.11 |
| Infec | NB | 0.988 | 1.22 |
| Renal | NB | 0.978 | 1.32 |
| Hepat | NB | 0.989 | 1.43 |
| Ear | NB | 0.977 | 1.21 |
| Preg | NB | 0.993 | 1.41 |
| Cong | NB | 0.995 | 1.88 |
| Neopl | NB | 0.998 | 1.22 |

Following pre-analysis, Poisson and ZIP models were fitted where Y = Endo and Immun, whereas NB and ZINB models were fitted where Y = Metab, Infec, Renal, Hepat, Ear, Preg, Cong and Neopl. Model performances were assessed by Akaike Information Criteria (AIC) and variance of Pearson residuals. AIC enabled the further calculation of weights of evidence.

### 3.4 Model performance results

Variance of Pearson residuals were displayed to provide a global overview of the fitting of all models. The reason why values of Pearson residuals were not shown here was because of the high proportion of zeroes present in the counts of the NB or Poisson distributed SOCs. Given that for all SOC counts, the proportions of zeroes were greater than 90%, mean Pearson residuals for all constructed models were estimated to have a range from −7.92E-06 to 0.032; if the absolute value of a Pearson residual is greater than 2 or 3 then it lacks statistical significance [29].

However, this does not take into account the rare nature of ADRs. Therefore, to circumvent this, the variance of all estimated Pearson residuals would serve as a more accurate measure of model fitting [30]. In the same situation where > 95% of the observed counts are zeroes, Lause et al. [31] recommended the variance of all estimated Pearson residuals as a diagnostic tool. In addition, they established if the variance of all Pearson residuals was close to 1, then the model captured the expected variability.

Therefore, in the context of our work, if the variance of all Pearson residuals was greater than 1.00, then a plausible signal may be produced. However, if the variance of all Pearson residuals was greater than the threshold of 1.00 by multiple folds, we argue that there may be overfitting or the model was unstable. As such, we have also included the AIC, ΔAIC and weight of evidence (w_Poisson, ZIP, ZINB or ZIP_) to detect potential agreement between multiple model diagnostics. Equations for AIC, ΔAIC and weight of evidence are found in [28].

For models where Y = Endo, Immun, Metab, Hepat, Ear and Preg, the corresponding ΔAIC values suggested that the ZI models provided a better fit than their non-ZI counterparts (NB vs ZINB, and Poisson vs ZIP). Among these, the NB and ZINB models where Y= Metab may be a signal of interest. This is because the variance of Pearson residuals for the NB and ZINB models were 1.15 and 1.18, respectively.

Secondly, ΔAIC and weight of evidence reflected that the ZINB model was the better supported model. Lastly, results shown in Table 6 and Figure 1 displayed agreement; Statistically significant ROR, PRR and IC for citalopram concerning Metab for females were observed (ROR, 2.27 [2.07, 2.48]; PRR, 2.21 [2.03, 2.41]; IC 1.13 [1.00, 1.26]).

**Table 6:**
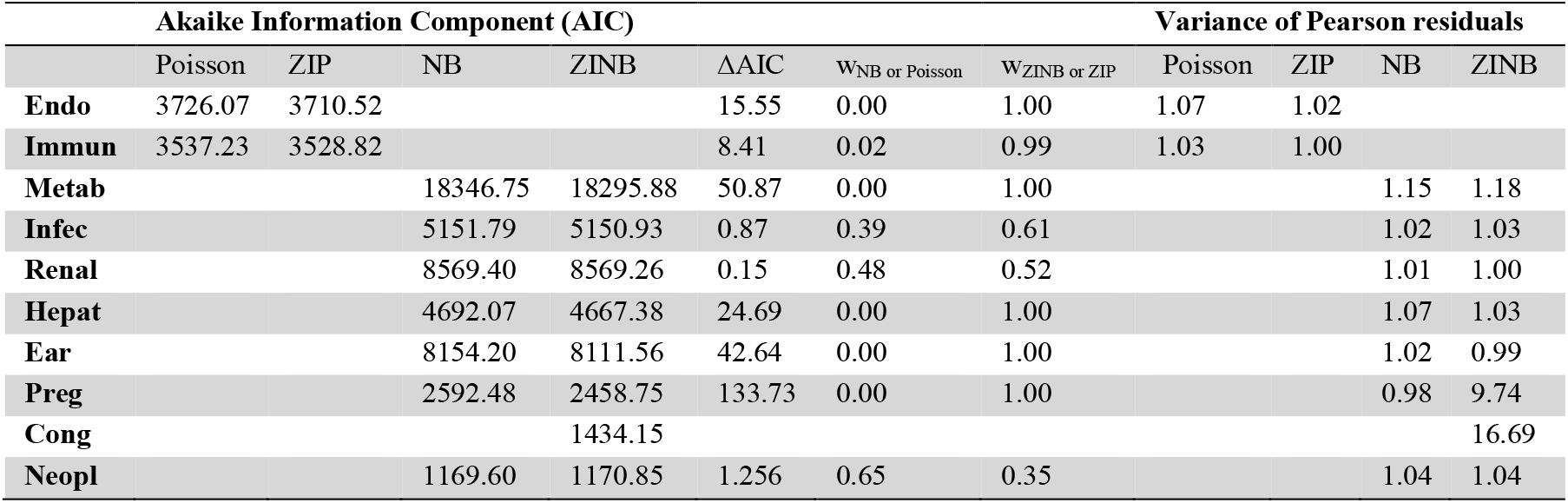
Mean Pearson residuals of positive AECs, AIC, delta AIC and weight. If ΔAIC> 10, then the model with the smaller AIC performed better. If ΔAIC= 4…7, then the model with the smaller AIC still performed better but with less support. If ΔAIC< 2, then the performance of both models are indistinguishable [27,28]. Only Poisson and ZIP models were fitted when Y = Endo or Immun; only NB or ZINB models were fitted when Y = Metab, Infec, Renal, Hepat, Ear, Preg, Neopl. The NB model failed to converge when Y = Cong, therefore weight of evidence could not be calculated.

**Fig 1:**
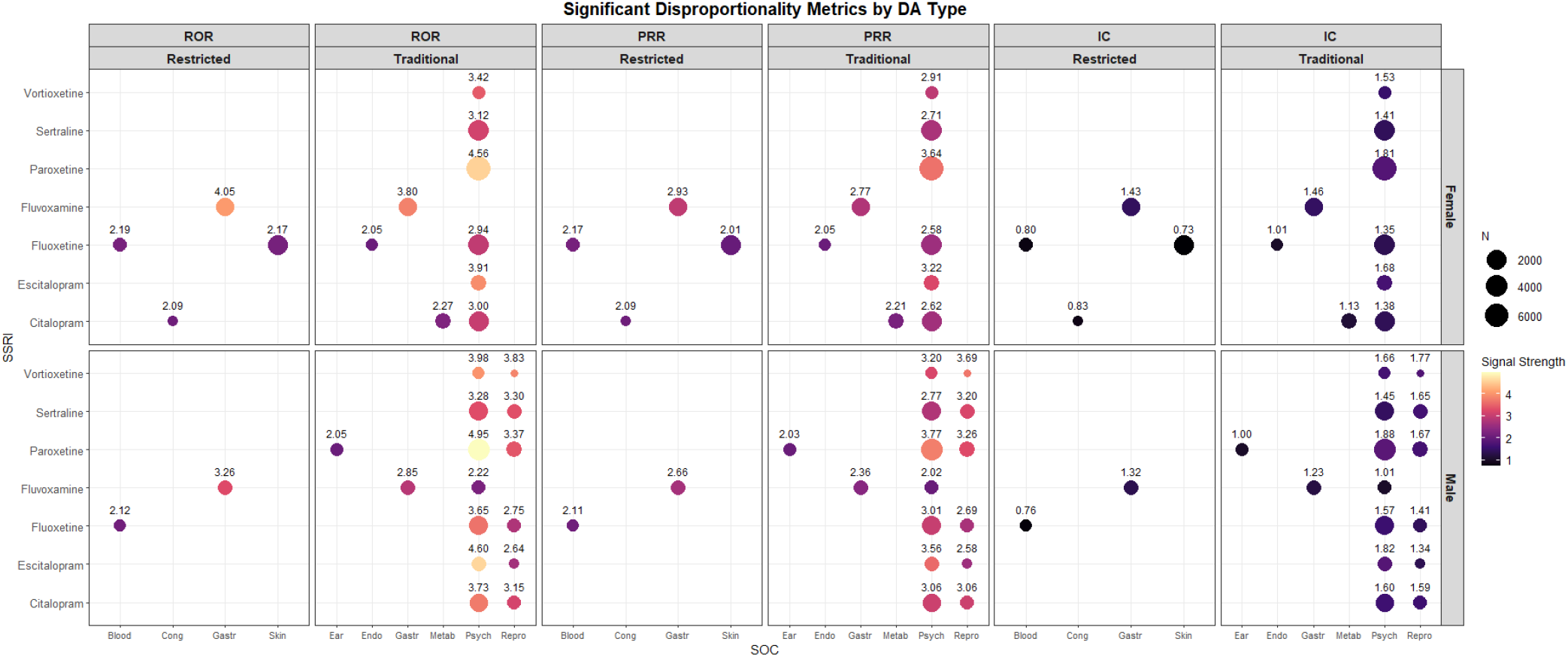
Dot plot summarising significant disproportionality metrics from traditional and restricted DA, stratified by sex. A ROR [95% CI] is significant when Lower 95% CI > 1.00; number of observed cases ≥ 5. A PRR [95% CI] is significant when PRR>2, lower 95% CI > 1.00; χ^2^≥ 4; number of observed cases ≥ 5. An IC [IC_025_, IC_0975_] is significant when IC_025_ is positive. The size of the dot is proportional to the number of cases. A different disproportionality metric is attributed to each third of this dot plot; from left to right, RORs, PRRs and ICs are shown, respectively. For each disproportionality metric, results of restricted DA are shown on the left, whereas results of the traditional DA are shown on the right. Female results are shown in the top row, male results are shown in the bottom row. The axis on the left displays SSRIs, axis at the bottom displays significant SOCs, axis on the right describes male and female). All significant results are displayed as forest plots and can be found in Supplementary Information (S8).

## Discussion

### 4.1 Justification for ZI models

A ZI model is a composite model comprised of a logistic component and a count component. Prior to model fitting, the researcher must establish the presence of two zero generating mechanisms (ZGMs), as the high proportion of zeroes in the response variable does not serve as viable motivation or an indicator for adopting ZI models [32]. In our work, we propose ZI models as a SDM as the first ZGM would generate structural zeroes due to ADRs which cannot occur; potentially because of physicochemical properties of the SSRI or biological implausibility. Simultaneously, we infer there is a second ZGM as an ADR may have occurred but remained unreported (underreporting).

At the beginning, we hypothesised that ADR data when transformed into counts may originate from a ZI data generating process (DGP). Given that for most of the ZI models, the AIC values were smaller than the non-ZI counterparts, and the weights of evidence of ZI models were also mostly greater in magnitude. This may support our hypothesis, even though AIC values of NB and ZINB models where Y=Neopl were 1169.60 and 1170.85, respectively.

Although this result may appear as disagreement with our hypothesis, this is not unexpected as during a simulation study by Feng where the aim was to assess the fit of count models to simulated data from Poisson, ZIP, NB and ZINB DGPs [33], a lower AIC for NB models when fitted to data from a ZINB DGP was estimated. Unsurprisingly, the majority of the AIC values of ZINB models were lower when fitted to data by a ZINB DGP. Thus, suggesting that during real-world application and specifically in the context of our work, if the observed ADR count data were to originate from a ZINB generating process, NB models may display lower AIC values than their ZI counterparts.

### 4.2 Use of an offset term

The first strength would be the use of an offset term (total number of reports of each SSRI) during model fitting. This approach draws parallels from DA, as reporting proportions for each SSRI would be modelled in place of absolute counts, as log(*µ*) = *β*_0_ + *β*_1_*X*_1_ + *⋯ β*_4_*X*_4_ + log(*offset*) can be rearranged such that *log* (*µ*⁄*offset*) = *β*_0_ + *β*_1_*X*_1_ + *⋯ β*_4_*X*_4_. This addresses model bias towards the most reported SSRI in the dataset.

### 4.3 Self-validation

The second strength would be the emphasis on self-validation. At the beginning of the workflow, SOCs were transformed into count data to prevent information loss. GoF tests were performed to determine NB or Poisson distribution within the SOC counts, and VMRs for overdispersion detection. During model fitting, firstly, models repeatedly underwent various assessments of absolute and relative model fit. The former was assessed by variance of Pearson residuals, obtained from estimated Pearson residuals; Pearsons’s residuals serve as a standardised measure of model fit given by *Pearson residual* 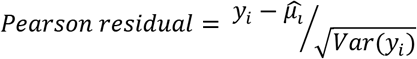.

Secondly, relative performance of models were compared by AICs, ΔAICs, and weights of evidence.

Lastly, the correction of *p*-values with the BH-method. The lack of *p*-value correction during multiple testing was a common limitation within PV studies. In addition to an overreliance on DA in PV studies, presenting a gap in SDMs which can be completed by the application of different statistical methods [34]. Two most commonly used *p*-value correction methods are the Bonferroni method and BH method [35]. The BH method was chosen as it is less conservative than the Bonferroni method, potentially increasing the false negative rate and rejecting true positive signals [35,36]. Both *p*-value correction methods have been used in *post-hoc* analyses in previous PV studies [37,38].

### 4.4 Potential utility of probability of structural zero (π)

The third strength would be the potential utility of the π. Whilst there has been work focussing on the application of ZI models in PV, the utility of the π remained unexplored. Firstly, the application of a zero-adjusted negative binomial (ZANB) to data of the Bulgarian PV database [39]; a ZANB model does not include a logistic component to model zeroes in the observed data whereas a ZINB model does. Secondly, the application of ZINB to simulated data and VAERS data [40]. However, the authors did not specify motivation for the need of a ZINB, nor did they present and discuss π [41]. We believe this to be a gap, as the π could contextualise outputs obtained from the count component. Given that underreporting is a characteristic flaw within SRSs, validation of predicted rates obtained from the count component would be of utility. Equally, the greater the π, we would interpret it as biological implausibility between a drug and a particular ADR.

### 4.5 Comparisons across all drugs within the same therapeutic class

The fourth strength would be the ability to compare across all drugs within the same therapeutic class, as well as individually between two drugs. Overall comparisons across all drugs were achieved by the application of the inverse link function to produce predicted values by age and stratified by male and female, whereas individual comparisons between two drugs would be represented by RR [95% CIs]. These results are considered significant when the BH corrected *p*-value < 0.05.

### 4.6 Use of SOCs

In this study there remains limitations. Firstly, designating Y as SOCs. We acknowledge that the use of PTs may produce results of greater accuracy than SOCs, however, SOCs were chosen as the Y of our models to reduce the number of tests conducted. The reason for this being correction decreases sensitivity, potentially increasing the likelihood of false negative results [42]. The hierarchical nature from SOC to PTs was also a point of consideration. As there are at least 11,000 PTs and 27 SOCs [43], designating each PT as an individual Y where each observation was a count may lead to an excessively high proportion of zeroes, preventing convergence of NB or ZINB models. There is preference for the analysis of PTs by regulatory bodies, however, given that the current objective was to display the operational capability of ZI models as a SDM, we believe this to be met. The potential implementation of our workflow at the PT level is an area which will be further explored.

### 4.7 Limited number of relevant variables

The number of biologically relevant variables were limited. In each iDAP, the only biologically relevant variables were age and gender. The absence of weight, BMI, height or smoking status as covariates during model construction prevented controlling of confounding by these variables, as well as a lack of variables describing comorbidities and polypharmacy. To optimise the future application of NB and ZINB models, datasets which contain these variables would be prioritised. Moreover, as the dataset extracted from the iDAPs of SSRIs was limited and simple. Stratification of model by gender and inclusion of interaction terms contributed to failed convergence. Only a fully adjusted model with variables shown in Table 2 converged. At our current stage, iDAP data may be considered a testbed and the fitting of NB and ZINB models has enabled the determination of what steps should be taken to control for confounders. Data extensiveness should also be a point of consideration as it could influence convergence of more complex or stratified models.

## Limitations

The Interactive Drug Analysis Profiles, from the MHRA, give a complete listing of all the spontaneous suspected ADRs reported through the Yellow Card scheme. While essential to safety monitoring, spontaneous reporting schemes have several inherent weaknesses. It is estimated that only 6% of ADRs are reported to regulatory authorities, which may lead to the underestimation of any given ADR. Under-reporting may vary by both reaction and by drug, even within the same class. Publicity about an adverse effect, length of time on the market, and novelty of the drug such as the first-in-class may also affect reporting. This can mean that comparisons between drugs using such reports can be problematic, particularly when small numbers are involved. Declines in reporting ADRs after the second year a drug has been on the market, known as the Weber effect, have been reported. Reporters are requested to report any suspected ADRs, and they do not have to demonstrate a clear causal link with the drug. This means that many reported ADRs may not be linked to the drug. Care needs to be taken with suspected fatal and other serious cases, where reporters may be more likely to err on the side of reporting due to the seriousness of the reaction. Confounding may also occur from previous exposure to SSRI (or related)-based therapy, or concomitant disease, either causing or contributing towards the reported ADR. Therefore, conclusions on the safety and risks of medicines cannot be made on the information obtained from the Drug Analysis Profiles alone. However, such data can be useful for hypothesis generation as in this statistical method development study.

## Conclusions

In conclusion, when SRS data is transformed into counts, or more specifically from SOCs to SOC counts, data may become samples from a ZI distribution-ZIP or ZINB. This enabled fitting of ZIP or ZINB models as potential SDMs, supplementing the current method of DA. At this current stage, we fitted these models to the SOC counts of iDAPs from YCS, where SSRIs were the drug class of interest. To ensure the robustness of signals, we suggest self-validation steps before and after model fitting, as important steps during this SGW. Prior to fitting of ZI models, GoF tests were conducted to determine which SOC counts were Poisson or NB distributed, and after model fitting, variance of Poisson residuals and weight of evidence from AIC were calculated. Future steps would be to potentially apply this SGW to another drug class, or when data permits, designating Y as deeper levels of the MedDRA hierarchy.

## Acknowledgements

We thank the MHRA for historical open-access data on ADRs.

## Statements and declarations

### Funding

The authors did not receive support from any organization for the submitted work.

### Prior postings and presentations, name(s) of any sponsor(s) of the research contained in the paper, along with grant number(s)

None to declare.

### Conflict of interest

Prof. Anthony Cox holds an honorary Pharmacovigilance Pharmacist position at the Yellow Card Centre West Midlands.

### Ethics approval

This project has been approved in line with the University of Birmingham’s research ethics processes (ERN_4560-Jun2025). Data used for analysis in this study is publicly available, fully anonymised data by the MHRA, an executive agency of the Department of Health and Social Care in the United Kingdom.

### Author contributions

All authors read and approved the final manuscript. Conceptualization – A.M.J., M.M.; Data curation – M.M.; Formal analysis – M.M., C.L.E., A.M.J., V.C., A.R.C.; Investigation – M.M.; Methodology – M.M., C.L.E.; Project administration – M.M., A.M.J.; Supervision – A.M.J., A.R.C., V.C.; Validation – M.M., C.L.E.; Visualization – M.M.; Writing – original draft – M.M.; Writing – review & editing – M.M., C.L.E., A.M.J., V.C., A.R.C.

### Data Availability statement

Additional data used in this study can be found in the accompanying supporting information and at: https://yellowcard.mhra.gov.uk/idaps and Man, L. Y. M. (Creator), Jones, A. M. (Supervisor) (16 Jun 2026). Research data supporting “Finding Safety Signals in Sparse Data: Zero-Inflated Models for Pharmacovigilance studies”. University of Birmingham. 10.25500/edata.bham.00001672

## Notes

### Author Declarations

This project has been approved in line with the University of Birmingham research ethics processes (ERN4560Jun2025). Data used for analysis in this study is publicly available, fully anonymised data by the MHRA, an executive agency of the Department of Health and Social Care in the United Kingdom.

