## Supplementary Information for "Finding Safety Signals in Sparse Data: Zero-Inflated Models for Pharmacovigilance studies"

S1: A systematic review by Jiao et al [15] which summarised the variety of SDMs deployed by various countries. Adapted from the review, this table presents SDMs where the number of countries which employ them is > 3.

| **Calculated metric** | **Category** | **Bayesian or Frequentist** | **Country** |
|---|---|---|---|
| Reporting Odds Ratio (**ROR**) | Disproportionality analysis | Frequentist | China, USA, UK, France, Japan, South Korea, Singapore, Thailand, Netherlands |
| Proportional Reporting Ratio (**PRR**) | Disproportionality analysis | Frequentist | China, USA, UK, France, Japan, Germany, South Korea |
| **I**nformation **C**omponent | Disproportionality analysis | Bayesian | Sweden, New Zealand, UK, France, China, USA, Japan, South Korea, Thailand, Singapore |
| Empirical Bayes Geometric Mean (**EBGM**) | Disproportionality analysis | Bayesian | USA, France, UK, China, Japan, Singapore, South Korea |
| Time to onset (**TTO**) **statistic** | Temporal / time-to-event | Frequentist | Netherlands, Canada, New Zealand, UK, Japan, China |
| **OR (LR)** | Regression | Frequentist | USA, China, Japan |

S2: Summary of the columns found in the *case, drug* and *event* CSV files.

| **case** | **drug** | **event** |
|---|---|---|
| *ADR* | *ADR* | *ADR* |
| *SEX* | *SEQ* | *SEQ* |
| *AGE_10* | *ROUTE* | *PT* |
| *RECVD_YEAR* | *MULTICONST* | *HLT* |
| *SENDER_TYPE* | *NONSERIOUS_SERIOUS_FATAL_NSF* | *HLGT* |
| *CONSUMER_YN* | | *SOC_ABBREV* |
| *HCP_YN* | | *FATAL_YN* |
| *NONSERIOUS_SERIOUS_FATAL_NSF* | | |

*ADR* sequential number identifying a single ADR report, *SEQ* sequential number identifying each adverse reaction within the ADR report *SEX* sex of patient as *Male, Female* or *Unknown*, *AGE_10* patient age at the time of the ADR expressed in 10-year bands with the lowest level of the band displayed in the file, *RECVD_YEAR* year report was first received by the MHRA, *SENDER_TYPE* describes the source of the ADR reports submitted to the MHRA, *CONSUMER_YN* Indicates if the ADR was reported by the consumer (the patient, their parent or a carer) of the product, *HCP_YN* Indicates if the ADR was reported by a healthcare professional, *NONSERIOUS_SERIOUS_FATAL_NSF* Indicates the seriousness of the ADR report, *MULTICONST* describes whether the drug in each ADR report contains more than one active ingredient, *PT* MedDRA Preferred Term, describing the adverse reaction, *HLT* MedDRA High Level Term which contains the corresponding Preferred Term, *HLGT* MedDRA High Level Group Term which contains the corresponding High Level Term, *SOC_ABBREV* MedDRA System Organ Class which contains the corresponding High Level Group Term

S3: 2x2 contingency table for disproportionality analysis (DA)

| | ADR of interest | Other ADRs |
|---|---|---|
| Drug of interest | a | b |
| All other drugs in the database or therapeutic class | c | d |

a= number of reports of drug of interest and ADR of interest; b = number of reports of drug of interest and other ADRs; c= number of reports of all other drugs in the database and ADR of interest; d= number of reports of all other drugs in the database and other ADR; when one conducts DA restricted to one therapeutic class, it is known as *restricted DA*. A summary of the advantages and disadvantages of traditional and restricted DA is presented by Cutroneo [44].

S4: Common disproportionality metrics and their respective uncertainty intervals, and thresholds for significance.

| **Disproportionality metric [Uncertainty interval]** | **Algorithm** | **Thresholds for significance** |
|---|---|---|
| **ROR [95% CI]** | $ROR = ad/bc$ | Lower 95% CI > 1.00; a ≥ 5 |
| **PRR [95% CI]** | $PRR = [a/(a+b)]/[c/(c+d)]$ | PRR>2, Lower 95% CI > 1.00; $\chi^2 \geq 4$; a≥ 5 |
| **IC [$IC_{025}$, $IC_{0975}$]** | $IC = log_2[\ [a(a+\cdots d)]/[(a+b)(a+c)]\ ]$ | $IC_{025} > 0.00$ |

ROR = Reporting Odds Ratio; PRR = Proportional Reporting Ratio; IC = Information Component; 95% CI = 95% confidence interval (Frequentist) or credibility interval (Bayesian); $IC_{025}$= lower limit (2.5$^{th}$ percentile) of IC credibility interval; $IC_{0975}$= upper limit (97.5$^{th}$ percentile) of IC credibility interval [44].

S5: Pre-analysis results of all 27 SOCs. This was conducted with the aim of determining the distribution of SOC counts.

| SOC | SOC (full description) | Variance-to-Mean ratio (VMR) | *p*-value (Poisson) | Follows Poisson | *p*-value (NB) | Follows NB |
|---|---|---|---|---|---|---|
| **Blood** | Blood and lymphatic system disorders | 1.2116 | 0 | No | 2.64E-09 | No |
| **Card** | Cardiac disorders | 1.302 | 0 | No | 2.04E-04 | No |
| **Cong** | Congenital, familial and genetic disorders | 1.8766 | 0 | No | 1.73E-01 | Yes |
| **Ear** | Ear and labyrinth disorders | 1.2131 | 0 | No | 2.14E-01 | Yes |
| **Endo** | Endocrine disorders | 1.038 | 1.97E-44 | No | 3.48E-01 | Yes |
| **Eye** | Eye disorders | 1.5522 | 0 | No | 7.96E-08 | No |
| **Gastr** | Gastrointestinal disorders | 1.4785 | 0 | No | 2.47E-31 | No |
| **Genrl** | General disorders and administration site conditions | 1.6468 | 0 | No | 0 | No |
| **Hepat** | Hepatobiliary disorders | 1.4264 | 0 | No | 2.89E-01 | Yes |
| **Immun** | Immune system disorders | 1.0096 | 3.60E-01 | Yes | 9.99E-01 | Yes |
| **Infec** | Infections and infestations | 1.2232 | 0 | No | 4.46E-02 | No |
| **Inj&P** | Injury, poisoning and procedural complications | 1.3553 | 0 | No | 1.20E-57 | No |
| **Inv** | Investigations | 1.5495 | 0 | No | 0 | No |
| **Metab** | Metabolism and nutrition disorders | 1.1077 | 0 | No | 1.73E-01 | Yes |
| **Musc** | Musculoskeletal and connective tissue disorders | 1.5163 | 0 | No | 8.85E-15 | No |
| **Neopl** | Neoplasms benign, malignant and unspecified (incl cysts and polyps) | 1.2177 | 0 | No | 2.57E-02 | No |
| **Nerv** | Nervous system disorders | 1.9331 | 0 | No | 0 | No |
| **Preg** | Pregnancy, puerperium and perinatal conditions | 1.4079 | 0 | No | 9.56E-01 | Yes |
| **Prod** | Product issues | 1.1657 | 0 | No | 3.92E-01 | Yes |
| **Psych** | Psychiatric disorders | 3.6964 | 0 | No | 1.80E-271 | No |
| **Renal** | Renal and urinary disorders | 1.3195 | 0 | No | 7.59E-01 | Yes |
| **Resp** | Respiratory, thoracic and mediastinal disorders | 1.4035 | 0 | No | 4.82E-05 | No |
| **Repro** | Reproductive system and breast disorders | 1.5 | 3.65E-150 | No | 8.78E-05 | No |
| **Skin** | Skin and subcutaneous tissue disorders | 1.2077 | 0 | No | 0 | No |
| **SocCi** | Social circumstances | 1.2941 | 0 | No | 9.96E-01 | Yes |
| **Surg** | Surgical and medical procedures | 1.1421 | 1.98E-95 | No | 7.63E-01 | Yes |
| **Vasc** | Vascular disorders | 1.1221 | 0 | No | 1.52E-04 | No |

S6: Descriptive statistics of the prepared dataset for DA with ‘pvda’. Descriptive statistics for traditional (left) and restricted (right) DA are both shown.

| | **All drugs in database (N= 3,098,534)** | | | **Only SSRIs (N= 112,285)** | | |
|---|---|---|---|---|---|---|
| **SOC** | Female Count (%) | Male Count (%) | Unknown Count (%) | Female Count (%) | Male Count (%) | Unknown Count (%) |
| **Blood** | 33,853 (47.68%) | 34,082 (48.00%) | 3062 (4.31%) | 519 (68.38%) | 218 (28.72%) | 22 (2.90%) |
| **Card** | 42,872 (50.33%) | 39,016 (45.80%) | 3299 (3.87%) | 1419 (66.84%) | 645 (30.38%) | 59 (2.78%) |
| **Cong** | 6566 (50.15%) | 3657 (27.93%) | 2870 (21.92%) | 131 (26.68%) | 156 (31.77%) | 204 (41.55%) |
| **Ear** | 11,883 (60.62%) | 7067 (36.05%) | 651 (3.32%) | 706 (65.67%) | 335 (31.16%) | 34 (3.16%) |
| **Endo** | 5214 (60.87%) | 2942 (34.35%) | 410 (4.79%) | 291 (75.78%) | 79 (20.57%) | 14 (3.65%) |
| **Eye** | 45,855 (60.66%) | 26,664 (35.27%) | 3076 (4.07%) | 1880 (71.95%) | 640 (24.49%) | 93 (3.56%) |
| **Gastr** | 237,318 (61.67%) | 132,669 (34.48%) | 14,806 (3.85%) | 10,082 (71.07%) | 3640 (25.66%) | 465 (3.28%) |
| **Genrl** | 223,142 (58.45%) | 138,651 (36.32%) | 19,956 (5.23%) | 8426 (66.81%) | 3717 (29.47%) | 468 (3.71%) |
| **Hepat** | 20,656 (52.28%) | 17,193 (43.51%) | 1665 (4.21%) | 311 (56.44%) | 225 (40.83%) | 15 (2.72%) |
| **Immun** | 26,733 (60.06%) | 15,644 (35.15%) | 2134 (4.79%) | 256 (74.85%) | 79 (23.10%) | 7 (2.05%) |
| **Infec** | 61,781 (51.72%) | 50,190 (42.02%) | 7482 (6.26%) | 400 (68.61%) | 162 (27.79%) | 21 (3.60%) |
| **Inj&P** | 68,879 (53.17%) | 49,288 (38.05%) | 11,379 (8.78%) | 1811 (62.02%) | 894 (30.62%) | 215 (7.36%) |
| **Inv** | 79,418 (46.28%) | 85,613 (49.89%) | 6574 (3.83%) | 1951 (61.94%) | 1056 (33.52%) | 143 (4.54%) |
| **Metab** | 35,845 (53.55%) | 27,514 (41.11%) | 3573 (5.34%) | 2063 (67.20%) | 873 (28.44%) | 134 (4.36%) |
| **Musc** | 75,510 (59.04%) | 47,564 (37.19%) | 4818 (3.77%) | 2235 (68.02%) | 946 (28.79%) | 105 (3.20%) |
| **Neopl** | 15,375 (49.62%) | 13,407 (43.27%) | 2205 (7.12%) | 66 (66.00%) | 30 (30.00%) | 4 (4.00%) |
| **Nerv** | 222,986 (60.30%) | 133,781 (36.18%) | 13,039 (3.53%) | 14,608 (67.34%) | 6340 (29.22%) | 746 (3.44%) |
| **Preg** | 17,788 (86.07%) | 1217 (5.89%) | 1661 (8.04%) | 309 (72.54%) | 48 (11.27%) | 69 (16.20%) |
| **Prod** | 12,202 (59.35%) | 6394 (31.10%) | 1964 (9.55%) | 177 (61.03%) | 80 (27.59%) | 33 (11.38%) |
| **Psych** | 130,820 (55.78%) | 95,815 (40.85%) | 7911 (3.37%) | 15,642 (61.78%) | 8759 (34.59%) | 918 (3.63%) |
| **Renal** | 24,741 (45.39%) | 27,171 (49.85%) | 2590 (4.75%) | 610 (55.40%) | 458 (41.60%) | 33 (3.00%) |
| **Repro** | 28,852 (62.84%) | 16,247 (35.39%) | 812 (1.77%) | 1198 (45.16%) | 1394 (52.54%) | 61 (2.30%) |
| **Resp** | 84,129 (58.35%) | 54,374 (37.71%) | 5685 (3.94%) | 1578 (67.75%) | 683 (29.33%) | 68 (2.92%) |
| **Skin** | 215,782 (62.93%) | 116,088 (33.86%) | 11,019 (3.21%) | 5947 (71.20%) | 2170 (25.98%) | 236 (2.83%) |
| **SocCi** | 2105 (46.99%) | 2093 (46.72%) | 282 (6.29%) | 143 (50.00%) | 131 (45.80%) | 12 (4.20%) |
| **Surg** | 6249 (51.07%) | 5004 (40.90%) | 982 (8.03%) | 70 (72.16%) | 20 (20.62%) | 7 (7.22%) |
| **Vasc** | 46,561 (59.14%) | 29,049 (36.90%) | 3120 (3.96%) | 972 (65.15%) | 468 (31.37%) | 52 (3.49%) |
| **Total** | 1,783,115 (57.55%) | 1,178,394 (38.03%) | 137,025 (4.42%) | 73,801 (65.73%) | 34,246 (30.50%) | 4,238 (3.77%) |

S7: Descriptive statistics of the prepared dataset for regression models (Poisson, NB, ZIP, ZINB). Variables which were not included in the final analysis are in bold.

| Variable | Type | Valid | Missing | Main Values / Notes |
|---|---|---|---|---|
| **SEX** | character | 41,127 | 0 | Female 68.1%, Male 31.9% |
| SEX_binary | Numeric | 41,127 | 0 | Female = 0: 68.1%,<br>Male = 1: 31.9% |
| AGE_10 | Numeric | 36,334 | 4,793 | 0: 231 (0.6%)<br>10: 1827 (5.0%)<br>20: 6234 (17.2%)<br>30: 7852 (21.6%)<br>40: 7103 (19.5%)<br>50: 4996 (13.8%)<br>60: 3466 (9.5%)<br>70: 2769 (7.6%)<br>80: 1611 (4.4%)<br>90: 245 (0.7%) |
| **HCP_YN** | Character | 41,127 | 0 | Y: 84%, N: 16% |
| HCP_YN_binary | Numeric | 41,127 | 0 | 1: 84%, 0: 16% |
| SSRI | Character | 41,127 | 0 | Citalopram: 6514 (15.8%)<br>Escitalopram: 1341 (3.3%)<br>Fluoxetine: 10885 (26.5%)<br>Fluvoxamine: 2460 (6.0%)<br>Paroxetine: 10638 (25.9%)<br>Sertraline: 8822 (21.5%)<br>Vortioxetine: 467 (1.1%) |

S8: Significant results shown as forest plots (ICs, PRRs, RORs). Full dataset of disproportionality metrics can be obtained upon request.

### Forest Plot of Significant ICs [95% CI]

| SSRI | N | IC | 95% CI | SOC |
|---|---|---|---|---|
| Traditional / Male | | | | |
| Paroxetine | 134 | 1.00 | [0.75, 1.24] | Ear |
| Fluvoxamine | 219 | 1.01 | [0.81, 1.19] | Psych |
| Fluvoxamine | 354 | 1.23 | [1.08, 1.38] | Gastr |
| Escitalopram | 44 | 1.34 | [0.88, 1.73] | Repro |
| Fluoxetine | 271 | 1.41 | [1.23, 1.57] | Repro |
| Sertraline | 1,578 | 1.45 | [1.38, 1.53] | Psych |
| Fluoxetine | 1,785 | 1.57 | [1.50, 1.64] | Psych |
| Citalopram | 241 | 1.59 | [1.41, 1.77] | Repro |
| Citalopram | 1,419 | 1.60 | [1.52, 1.67] | Psych |
| Sertraline | 308 | 1.65 | [1.49, 1.81] | Repro |
| Vortioxetine | 82 | 1.66 | [1.33, 1.95] | Psych |
| Paroxetine | 489 | 1.67 | [1.54, 1.80] | Repro |
| Vortioxetine | 16 | 1.77 | [0.98, 2.39] | Repro |
| Escitalopram | 358 | 1.82 | [1.67, 1.97] | Psych |
| Paroxetine | 3,314 | 1.88 | [1.83, 1.92] | Psych |
| Restricted / Male | | | | |
| Fluoxetine | 80 | 0.76 | [0.43, 1.06] | Blood |
| Fluvoxamine | 354 | 1.32 | [1.16, 1.46] | Gastr |
| Traditional / Female | | | | |
| Fluoxetine | 92 | 1.01 | [0.70, 1.29] | Endo |
| Citalopram | 500 | 1.13 | [1.00, 1.26] | Metab |
| Fluoxetine | 2,893 | 1.35 | [1.29, 1.40] | Psych |
| Citalopram | 2,156 | 1.38 | [1.31, 1.44] | Psych |
| Sertraline | 3,094 | 1.41 | [1.36, 1.46] | Psych |
| Fluvoxamine | 1,247 | 1.46 | [1.38, 1.54] | Gastr |
| Vortioxetine | 152 | 1.53 | [1.29, 1.75] | Psych |
| Escitalopram | 556 | 1.68 | [1.56, 1.80] | Psych |
| Paroxetine | 6,363 | 1.81 | [1.78, 1.85] | Psych |
| Restricted / Female | | | | |
| Fluoxetine | 2,073 | 0.73 | [0.67, 0.79] | Skin |
| Fluoxetine | 190 | 0.80 | [0.59, 1.00] | Blood |
| Citalopram | 36 | 0.83 | [0.32, 1.26] | Cong |
| Fluvoxamine | 1,247 | 1.43 | [1.35, 1.51] | Gastr |

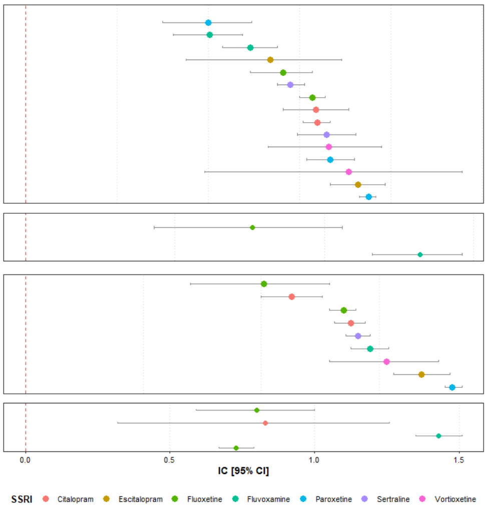


### Forest Plot of Significant PRRs [95% CI]

| SSRI | N | PRR | 95% CI | SOC |
|---|---|---|---|---|
| Traditional / Male | | | | |
| Fluvoxamine | 219 | 2.02 | [1.79, 2.28] | Psych |
| Paroxetine | 134 | 2.03 | [1.72, 2.41] | Ear |
| Fluvoxamine | 354 | 2.36 | [2.16, 2.58] | Gastr |
| Escitalopram | 44 | 2.58 | [1.93, 3.45] | Repro |
| Fluoxetine | 271 | 2.69 | [2.39, 3.02] | Repro |
| Sertraline | 1,578 | 2.77 | [2.65, 2.90] | Psych |
| Fluoxetine | 1,785 | 3.01 | [2.89, 3.13] | Psych |
| Citalopram | 1,419 | 3.06 | [2.92, 3.20] | Psych |
| Citalopram | 241 | 3.06 | [2.70, 3.47] | Repro |
| Sertraline | 308 | 3.20 | [2.86, 3.57] | Repro |
| Vortioxetine | 82 | 3.20 | [2.66, 3.86] | Psych |
| Paroxetine | 489 | 3.26 | [2.99, 3.56] | Repro |
| Escitalopram | 358 | 3.56 | [3.26, 3.89] | Psych |
| Vortioxetine | 16 | 3.69 | [2.29, 5.94] | Repro |
| Paroxetine | 3,314 | 3.77 | [3.66, 3.88] | Psych |
| Restricted / Male | | | | |
| Fluoxetine | 80 | 2.11 | [1.60, 2.77] | Blood |
| Fluvoxamine | 354 | 2.66 | [2.42, 2.92] | Gastr |
| Traditional / Female | | | | |
| Fluoxetine | 92 | 2.05 | [1.67, 2.52] | Endo |
| Citalopram | 500 | 2.21 | [2.03, 2.41] | Metab |
| Fluoxetine | 2,893 | 2.58 | [2.49, 2.67] | Psych |
| Citalopram | 2,156 | 2.62 | [2.52, 2.72] | Psych |
| Sertraline | 3,094 | 2.71 | [2.62, 2.79] | Psych |
| Fluvoxamine | 1,247 | 2.77 | [2.65, 2.90] | Gastr |
| Vortioxetine | 152 | 2.91 | [2.53, 3.35] | Psych |
| Escitalopram | 556 | 3.22 | [3.00, 3.47] | Psych |
| Paroxetine | 6,363 | 3.64 | [3.56, 3.72] | Psych |
| Restricted / Female | | | | |
| Fluoxetine | 2,073 | 2.01 | [1.91, 2.12] | Skin |
| Citalopram | 36 | 2.09 | [1.43, 3.07] | Cong |
| Fluoxetine | 190 | 2.17 | [1.82, 2.59] | Blood |
| Fluvoxamine | 1,247 | 2.93 | [2.79, 3.07] | Gastr |

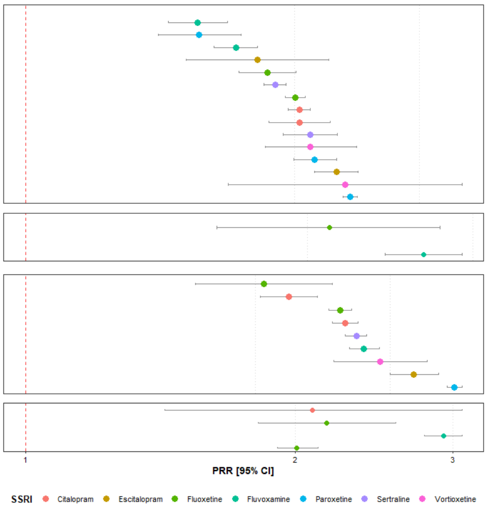


### Forest Plot of Significant RORs [95% CI]

| SSRI | N | ROR | 95% CI | SOC |
|---|---|---|---|---|
| Traditional / Male | | | | |
| Paroxetine | 134 | 2.05 | [1.72, 2.43] | Ear |
| Fluvoxamine | 219 | 2.22 | [1.92, 2.57] | Psych |
| Escitalopram | 44 | 2.64 | [1.95, 3.57] | Repro |
| Fluoxetine | 271 | 2.75 | [2.44, 3.11] | Repro |
| Fluvoxamine | 354 | 2.85 | [2.53, 3.22] | Gastr |
| Citalopram | 241 | 3.15 | [2.77, 3.59] | Repro |
| Sertraline | 1,578 | 3.28 | [3.10, 3.47] | Psych |
| Sertraline | 308 | 3.30 | [2.94, 3.70] | Repro |
| Paroxetine | 489 | 3.37 | [3.07, 3.69] | Repro |
| Fluoxetine | 1,785 | 3.65 | [3.46, 3.85] | Psych |
| Citalopram | 1,419 | 3.73 | [3.51, 3.96] | Psych |
| Vortioxetine | 16 | 3.83 | [2.32, 6.34] | Repro |
| Vortioxetine | 82 | 3.98 | [3.09, 5.12] | Psych |
| Escitalopram | 358 | 4.60 | [4.07, 5.21] | Psych |
| Paroxetine | 3,314 | 4.95 | [4.75, 5.15] | Psych |
| Restricted / Male | | | | |
| Fluoxetine | 80 | 2.12 | [1.61, 2.79] | Blood |
| Fluvoxamine | 354 | 3.26 | [2.87, 3.70] | Gastr |
| Traditional / Female | | | | |
| Fluoxetine | 92 | 2.05 | [1.67, 2.53] | Endo |
| Citalopram | 500 | 2.27 | [2.07, 2.48] | Metab |
| Fluoxetine | 2,893 | 2.94 | [2.82, 3.06] | Psych |
| Citalopram | 2,156 | 3.00 | [2.86, 3.15] | Psych |
| Sertraline | 3,094 | 3.12 | [3.00, 3.25] | Psych |
| Vortioxetine | 152 | 3.42 | [2.86, 4.10] | Psych |
| Fluvoxamine | 1,247 | 3.80 | [3.54, 4.08] | Gastr |
| Escitalopram | 556 | 3.91 | [3.55, 4.30] | Psych |
| Paroxetine | 6,363 | 4.56 | [4.43, 4.69] | Psych |
| Restricted / Female | | | | |
| Citalopram | 36 | 2.09 | [1.43, 3.07] | Cong |
| Fluoxetine | 2,073 | 2.17 | [2.05, 2.30] | Skin |
| Fluoxetine | 190 | 2.19 | [1.83, 2.62] | Blood |
| Fluvoxamine | 1,247 | 4.05 | [3.77, 4.36] | Gastr |

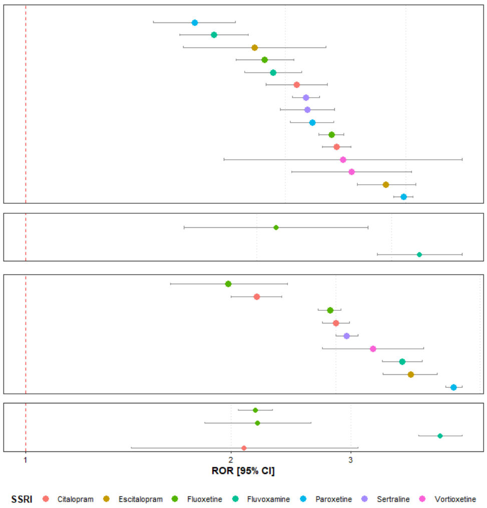
